# Predictive Capacity of COVID-19 Test Positivity Rate

**DOI:** 10.1101/2021.03.04.21252897

**Authors:** Livio Fenga, Mauro Gaspari

## Abstract

COVID-19 infections can spread silently, due to the simultaneous presence of significant numbers of both critical and asymptomatic to mild cases. While for the former reliable data are available (in the form of number of hospitalization and/or beds in intensive care units), this is not the case of the latter. Hence, analytical tools designed to generate reliable forecast and future scenarios, should be implemented to help decision makers planning ahead (e.g. medical structures and equipment). Previous work of one of the authors shows that an alternative formulation of the Test Positivity Rate (TPR), i.e. the proportion of the number of persons tested positive in a given day, exhibits a strong correlation with the number of patients admitted in hospital and intensive care units. In this paper, we investigate the lagged correlation structure between the newly defined TPR and the hospitalized people time series, exploiting a rigorous statistical model, the Seasonal Auto Regressive Moving Average (*SARIMA*). The rigorous analytical framework chosen, i.e. the stochastic processes theory, allowed for a reliable forecasting about 12 days ahead, of those quantities. The proposed approach would also allow decision makers to forecast the number of beds in hospitals and intensive care units needed 12 days ahead. The obtained results show that a standardized TPR index is a valuable metric to monitor the growth of the COVID-19 epidemic. The index can be computed on daily basis and it is probably one of the best forecasting tools available today for predicting hospital and intensive care units overload, being an optimal compromise between simplicity of calculation and accuracy.

## 1 Introduction

One of the aspects that makes the COVID-19 pandemic difficult to control, is the simultaneous presence of significant numbers of both critical and asymptomatic to mild cases. While for the former reliable data are available (in the form of number of hospitalizations and/or beds in ICUs), this is not the case of the latter [35, 13, 18]. In many instances, in fact, those who contracted the virus are unaware of such a condition and thus enter the status of spreaders or, in the worse case, super-spreader. Such a phenomenon, commonly referred to as under-ascertainment, is the primary reason for a disease to spread uncontrolled. Should it be not carefully checked nor effectively counteracted, it can potentially grow indefinitely, posing severe health problems at a global level and severely impacting whole health systems. Action-wise, such a situation calls for at least two measures: on the one hand policy and decision makers should plan ahead the needs in terms of medical structures and equipment whereas, on the other hand, analytical tools designed to generate reliable forecast and future scenarios should be implemented. While a number of effective approaches have been studied and proposed for different epidemics over the years, this is not the case of the CoVID-19 pandemic. In fact, all the efforts so far done to model and predict such a disease might hardly support the idea that a uniformly “better” model is available to describe and predict the evolution of such a catastrophic pandemic. Therefore, even though many valid contributions have been proposed so far [24], it is not unreasonable to look at those efforts as the building block of one or more best practices. In particular, the forecasting problem has been addressed for two of the the most populated countries in the world, i.e. China [26] and India [37]. A survey including other approaches is presented here [38]. The complexity of such a task is discussed in [4], where the authors analyzed three different regional-scale models for forecasting and assessing the course of the pandemic. Along those lines, is worth mentioning the excellent article [23], where the main reasons leading to the failure of a forecasting models are presented. Finally, two different predictive approaches has been proposed for Italy, i.e. one exploiting the bootstrapped prediction generated by a model of the type ARMA [14] and one based on the simulated annealing algorithm [15].

The Test Positivity Rate (TPR) is one of the indexes often used worldwide for monitoring the progression of the COVID-19 pandemic, see for example the coronavirus testing dataset [20], which contains an updated picture of the international situation concerning testing strategies and the associated data for many countries. Until now, the TPR was mainly studied considering its relationship with confirmed cases [12], for example it was used to estimate COVID-19 prevalence in the different states of US [30]. However, a more intensive use of diagnosis tests associated with a standardization of the TPR, crucial in light of differences in the available tests, can solve their limited investigation abilities (see, e.g., [32]).

In more details, a recent work of one of the authors [19] shows that a standardized COVID-19 Test Positivity Rate (TPR) can be used to predict hospital overload. In particular, by observing its trend, it is possible to forecast the course of patients admitted in hospital and in intensive care units. For example, when the TPR reaches a peak, a growth in COVID-19 hospitalisations lasting 12-15 days can be inferred.

There is an intuitive motivation behind such a behavior: COVID-19 epidemiological data show that symptoms, on average, occur 11 days after the contraction of the infection and that critical patients are admitted in hospital about 4 days later. If we assume that the TPR is a measurement of the infections occurring in a given day, in an ideal situation, the infected people with a critical evolution will be presumably admitted in hospital 15 days later. More precisely, if the TPR increases in a given day, an increasing number of active cases (including the unknown ones) can be inferred for the same day, and presumably the number of infections is increasing too. Thus, after a while the number of hospitalized people will also increase. In other words, the insight is that the TPR index models the trend of the COVID-19 infections, and it is designed to embody the unknown cases. Clearly, for this measure to be valid, all the administered diagnostic tests should be considered in the TPR calculation, as pointed out in [19]. However, there are known biases involving diagnostic tests data that are difficult to deal with, e.g., those related to reporting delays [20]. As a result, the ideal predictive capacity cannot be assumed in practice, especially if different kind of tests are used, as in the case of the current Italian situation.

Despite these limitations, the TPR can be effectively used to deduct important information on the course of the disease, as illustrated in Figure 1, where the epidemic course in Toscana region in autumn 2020 is depicted. This Figure also plots the time series of patients admitted in hospitals and in intensive care units. An interesting correlation between the curves can be observed: the TPR peak anticipates the peak of patients admitted in hospital and intensive care units.

**Figure 1:**
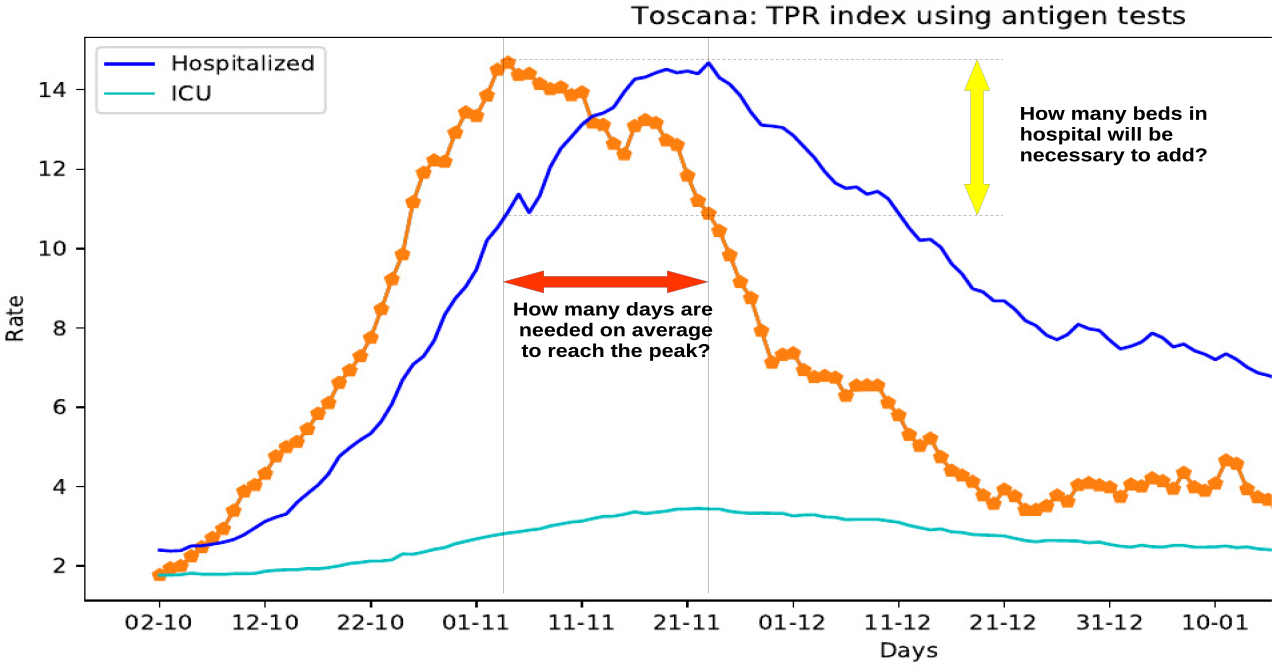
The TPR index predictive capacity.

The aim of this research is to analyse in details this scenario to get to the heart of some hard-hitting questions, especially when the TPR is growing considerably. How many days will be needed to reach the peak of hospitalized people? How many beds in hospitals will be necessary to add? And, in general, which is the “theoretical” predictive capacity of the proposed TPR index?

Starting from this motivation, we analysed the TPR index time series, as well as the hospitalized, and ICU patients time series, to investigate the predictive capacity of the TPR index, e.g, to individuate the time lags that can be effectively inferred from the available data. We first introduce the statistical methodology used and then we present a detailed analysis for four Italian region, for which data on antigen tests were available as reported in [19].

The lagged correlation between the TPR and hospitalized people time series will be modeled using a rigorous statistical model, i.e. of the type *SARIMA* (short for Seasonal Auto Regressive Moving Average). A detailed description of the underlying mathematics is presented in the Methods section.

A generalization of the *ARIMA* (Auto Regressive Moving Average) class [7], *SARIMA* models have been introduced to model complex dynamics of the type stochastic seasonal in many fields of research, such as economics [16] and [11], engineering [28] or hydrology [29]. In epidemiology, *SARIMA* models have been applied in a variety of studies: in [31] the authors applied this model for estimating case occurrence of two diseases: malaria and hepatitis A from January 1980 to June 1995 for the United States whereas in [10] the epidemiological and aetiological characteristics of influenza have been identified by establishing suitable SARIMA models. In particular, such an approach proved to be accurate in the forecasting of the percentage of visits for influenza-like illness in urban and rural areas of Shenyang (China). More recently, [27] used the SARIMA method – in conjunction with models belonging to the class exponential smoothing – to predict the trend of acute hemorrhagic conjunctivitis disease and used the obtained outcomes to provide evidence for the government to formulate policies regarding its prevention in mainland China.

The proposed mathematical model allowed us to estimate a predictive lag of about 12 days of the TPR for the prediction of hospitalized people time series in some Italian regions. Moreover, we defined a methodology to forecast the number of beds in hospitals and intensive care units needed 12 days ahead. The obtained results show that a standardized TPR index is a valuable metrics to monitor the growth of the COVID-19 epidemic. The index can be computed daily and it is probably one of the best forecasting tools available today for monitoring hospital and intensive care units overload, being an optimal compromise between simplicity of calculation and accuracy.

## 2 Results

The data used in this paper are made available by the Italian Civil Protection Department and publicly accessible, free of charge, at the following web address: https://github.com/pcm-dpc. In more details, these data – sampled at a daily frequency – are those necessary to compute the TPR (the number of new persons tested positive for COVID-19; the number of tests done considering both molecular (PCR) tests and antigen tests, and the number of healed persons), and those related to the number of hospitalizations and beds in intensive care units occupied by patients tested positive for COVID-19. The considered time frame ranges from Sept. 2 2020 to Feb. 10 2021 for a total of 353 data points. We have analysed 4 Italian regions for which the collection of the data on the antigen-based tests administered from Oct. 2020 to the 15th of Jan. 2021, has been possible, i.e. Toscana, Veneto, Piemonte and Alto Adige. The interested reader may refer to [19] for the details of the data collection procedure. Unfortunately, certain data concerning the use of diagnosis tests in the considered time frame are still not available for the other Italian regions. Figure 2 presents the TPR and hospitalised time series for Toscana, Veneto, Piemonte and Alto Adige.

**Figure 2:**
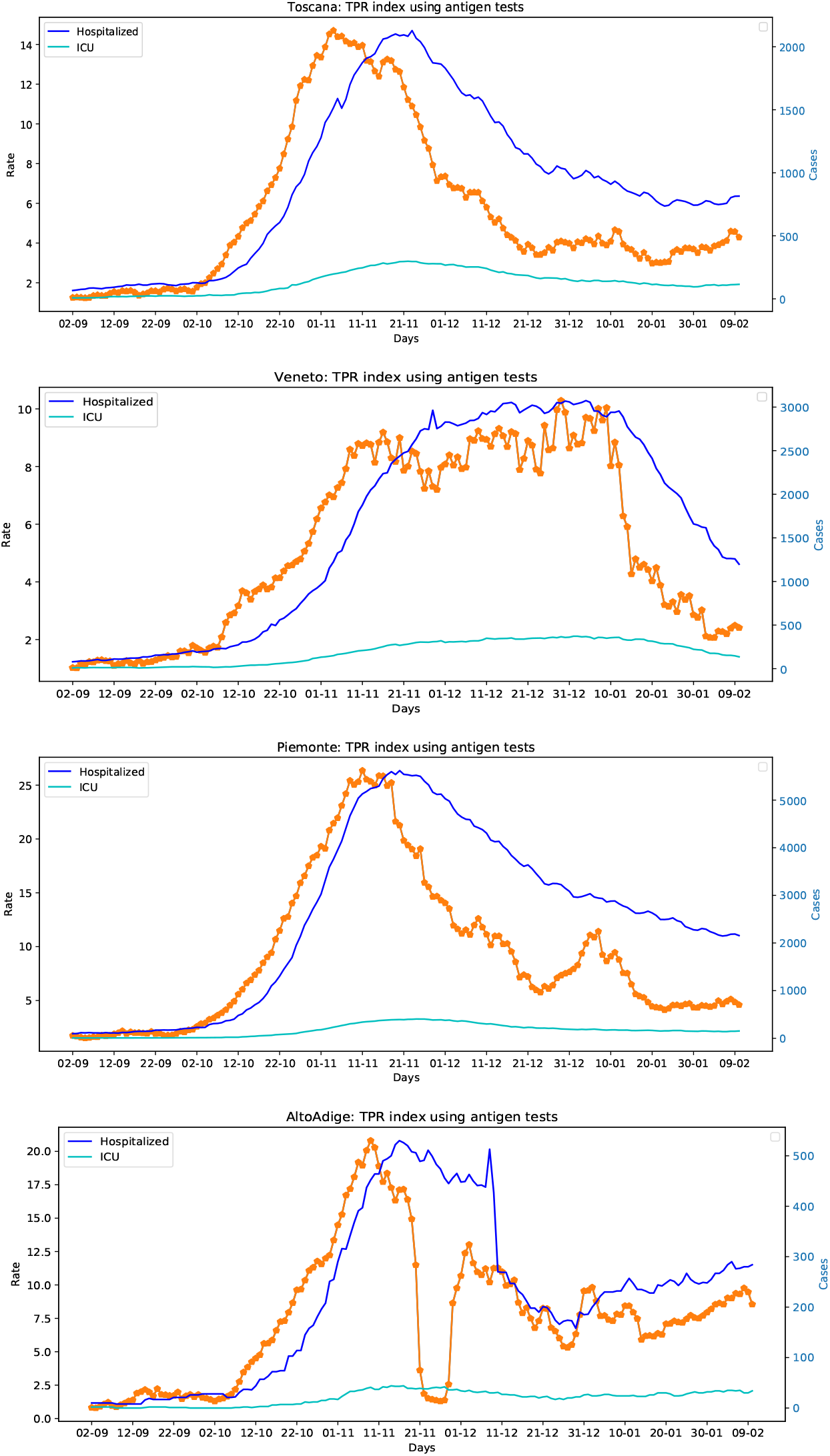
The TPR index and hospitalized patients time series of Toscana, Veneto, Piemonte and Alto Adige.

The presented empirical experiment considers two different scenarios, according to the way the available information is used. Their aim is to answer the hard-hitting questions that we have set in Figure 1. The first one – which can be defined of the type *real-life* – exploits the whole data set and it is designed to analyse the predictive capacity of TPR, to deliver a “theoretical” time lag between the two series, and prediction which, by design, cannot be verified being projected into the unknown future. On the contrary, the second experiment concerns forecasting the number of beds needed in hospitals and intensive care units after the determined time lag in specific situations in the past, that can be verified using the available data.

### 2.1 Analysis of the TPR predictive capacity

In essence, this part of the experiment, being based on the whole data set, can support only qualitative considerations on the proposed method. In accordance with the intuition that TPR represents the evolution of infections, the TPR should impact the hospitalization time series 15 days in advance. Studying the lagged correlations between the TPR time series and those of patients admitted in hospitals and ICUs, using the SARIMA model, we have individuated a predictive time lag of about 12 days for all the analysed regions, which confirm our intuitive hypothesis. Indeed, a 12 days predictive capacity for the TPR, with respect to hospitalized patients instead of the hypothesised 15, can be reasonably expected considering the above mentioned retrospective revisions effect [20]. In Table 1, we will report the time lag estimated for each region, along with an approximated multiplier accounting for the positive (negative) variation in the number of beds needed for a unit increase (decrease) of the TPR index.

**Table 1:** This table presents the results of the regression models with SARIMA errors concerning patients admitted in hospitals and intensive care units for Toscana, Veneto, Alto Adige and Piemonte regions. The columns *Days* and *Beds* indicate the TPR predictive capacity in days (with the associated t-value) and the estimated variation of beds in both hospitals and ICUs.

| Region | Hospitalized |  |  | ICU |  |  |
| --- | --- | --- | --- | --- | --- | --- |
|  | Days | t-value | Beds | Days | t-value | Beds |
| Toscana | 12 | 2.34 | 54 | 12 | 2.05 | 9 |
| Piemonte | 12 | 3.82 | 86 | 12 | 2.03 | 36 |
| Veneto | 13 | 2.07 | 82 | 13 | 2.52 | 12 |
| Alto Adige | 12 | 1.92 | 30 | 12 | 5.60 | 8 |

As estimates of future values yet to realize, these predictions can be mainly exploited to make qualitative inferences. For example, in the Veneto region, if the TPR increases of one unit, the model estimates that 82 additional beds may be needed in the near future (after 12 days). As for the ICUs we can expect 12 additional beds. Vice versa, if the TPR decreases in Veneto a similar amount of beds should be subtracted. In the considered regions, the average variation of beds in hospital and ICUs are 63 and 16 respectively.

### 2.2 Forecasting hospital overload

The second scenario envisioned, has been designed to carry out a precise evaluation of the performances delivered by the proposed method. To do so, we employed a test set with the same length but different starting point, as illustrated in Table 2. In practice, both structure and parameters of each SARIMA models has been estimated on the training set (this time with different sample sizes but same starting point) and, as already mentioned, evaluated on a “unknown” portion of data. Such a quantitative evaluation has been conducted considering different scenarios on all the studied regions: two in which the TPR was growing considerably in Toscana and Alto Adige; one associated to the beginning of the “red zone”^1^ Piemonte; one characterized by a slow growth of the TPR index in Veneto; and one associated to a fast lowering of the TPR indicator in Veneto.

**Table 2:** Forecasting dates in different situations: training and test set

| Region | Situation | Training set | Obs | Test set | Obs |
| --- | --- | --- | --- | --- | --- |
| Toscana | fast growing | 02/09/20 – 10/31/20 | 60 | 01/11/20 – 15/11/20 | 15 |
| Piemonte | Red Zone start | 02/09/20 – 06/11/20 | 66 | 07/11/20 – 22/11/20 | 15 |
| Veneto | slow growing | 02/09/20 – 09/12/20 | 99 | 13/12/20 – 28/12/20 | 15 |
| Veneto | fast lowering | 02/09/20 – 29/12/20 | 136 | 15/01/21 – 30/01/21 | 15 |
| Alto Adige | fast growing | 02/09/20 – 04/11/20 | 64 | 05/11/20 – 20/11/20 | 15 |

As for the REG-SARIMA model, as described in the Methods section, the model order has been defined using the MAICE procedure and constraining the Box-Cox *λ* parameter to 0 (i.e. log – transforming the data). However, being an exhaustive search of the “best” REG-SARIMA model either unfeasible or or impractical for computational reasons, the competition set has been built following the Box-Jenkins procedure, as illustrated, e.g., in [7]. Almost all the parameters of the final models are statistically significant and generate a sequence of residuals which can be deemed acceptable in terms of whiteness. Most of the times, the Maximum Likelihood algorithm converged quickly, with the only exception of the Piemonte region. In this case, a “sparse” data generating process in the autoregressive part involved a lengthy estimation approach – of the type trial and error – for the definition of the “best” (in AIC sense) model’s non-seasonal structure.

As already pointed out, the adopted MAICE procedure 13 is constrained to a specific value of the Box-Cox constant, which therefore has been set to *λ*_0_ = 0. As for the maximum order Γ_0_, it has been arbitrarily chosen on a case by case basis (see the Methods section for details).

The results of the forecasting experiments are summarized in Figure 3. The reader will certainly notice that the best forecasting results are obtained in the last experiment concerning the fast TPR lowering scenario in the Veneto region, where more data are available. However, all the other example provide reasonable results, and most importantly, when a fast growing of the TPR was present in the preceding of the cut, significant increases in hospitalizations are estimated. The determined increments are in general comparable to the generic estimations presented in Table 1.

**Figure 3:**
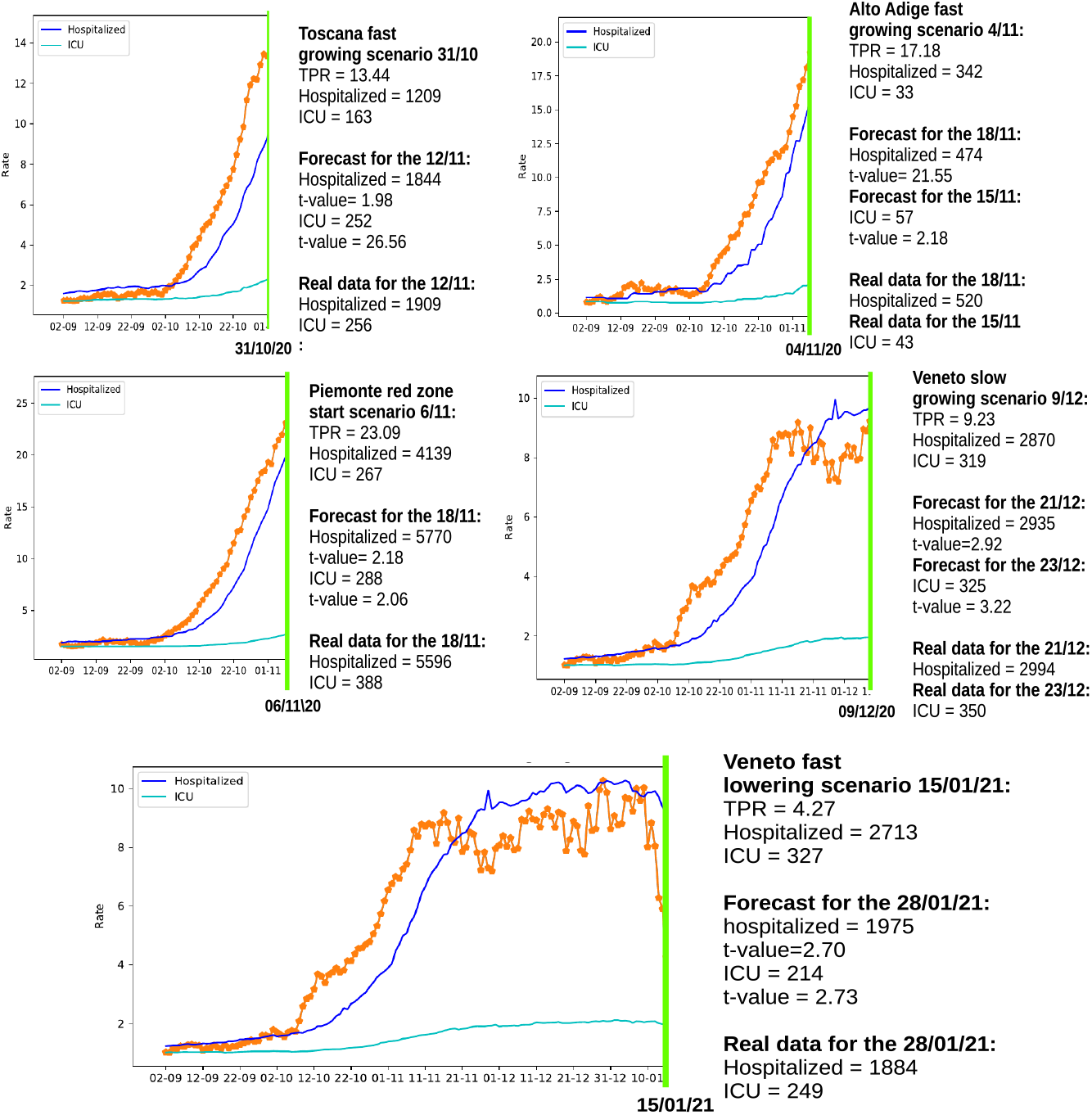
Forecasting hospitalized patients growth in 5 different scenarios for regions: Toscana, Alto Adige, Piemonte and Veneto (also including a fast lowering example).

## 3 Discussion

The proposed approach is general and can be exploited in any region/state under the condition that a set of requirements, below reported, are satisfied:

1. The data on the antigen tests administrated are provided;
2. The time series of new positive cases should include the daily number of new positives tested using only antigen tests;
3. The TPR should reach a peak before the hospitalized and ICU patients reach theirs.

The third criterion captures the same effect dealt with in [35, 18]. In particular in [35] it is stated that “the peak of the cases curves shifts when they are adjusted for under-ascertainment”. The rationale behind this idea is that the peak of unknown infections necessarily precedes the one related to the hospital admissions.

In general, when the first two requirements hold, then the 3rd one should hold as well. Vice-versa, if this is not the case, probably other anomalies or errors occur in the data. Moreover, issues concerning tests reliability cannot be excluded a priori – expecially when the ratio between hospitalized and positive cases growth considerably (e.g. due to tests specificity issues which might be related to new variants [3]). Should one or more of the above mentioned requirements be unfulfilled, the predictive properties of TPR might be affected. If this the case, an integration effort should be made to collect the missing data, and/or correct possible errors. For example, even though requirement 2 was not met for the Alto Adige region, we were able to analyse the TPR by manually adding the missing information to the time series of the new positives [19].

At this point, it is worth to compare the TPR index with other COVID-19 key indicators, commonly used for monitoring purposes [17], to the end of assessing their predictive properties. In particular, we have chosen the following indicators, designed to measure the dynamical behavior of the infections, i.e:

- *Growth rate*: positives daily variation;
- *Incidence*: fraction of COVID-19 positives per 100.000 individuals;
- *The reproduction number R*_*t*_: number of secondary infections generated from a case at time t.

Table 3 shows the pure predictive capacity with respect to hospitalizations of these COVID-19 indicators, for comparison with the TPR. While the TPR can be considered as a measure of the number of infections that occur on a certain day, also accounting for unknown cases, indicators based on officially reported positive cases (e.g. incidence and growth rate), measure the variation of official cases in a given area. Assuming that critical cases are admitted into hospitals within 4 days after tested positive, such a delay can be taken as a approximate “upper bound” for their pure predictive capacity.

**Table 3:** Pure predictive capacity in days of different COVID-19 indicators with respect to hospitalization.

| Metrics | What it represents | Days |
| --- | --- | --- |
| TPR | Number of active cases in a region also embodying the unknown portion of asymptomatic | 15 |
| Growth rate | Variation of detected positive cases in a region | 4 |
| Incidence | Number of known cases in a region | 4 |
| $R_t$ index | Variation of the infections dynamics in a region | 4 |

**Table 4:** This table presents the detailed results of the experiment presented in Section 2.1, for studying the SARIMA lagged correlation between the TPR time series and those of patients admitted in hospitals and ICUs. The last two columns *Days* and *Beds* indicate the TPR predictive capacity in days and the number of additional beds in hospital or ICU after 12 days for each TPR unit.

|  |  |  |  |  |  |  |  |  |  |  |
| --- | --- | --- | --- | --- | --- | --- | --- | --- | --- | --- |
| **Toscana** |  |  |  |  |  |  |  |  |  |  |
| SARIMA(2,1,0)(1,0,1) Box Cox trans: $\lambda=1.2$ $\beta_{t-value}=2.34$ | | | | | | | | | | |
| | $\phi_1$ | $\phi_2$ | $\Phi_1$ | $\Theta_1$ | $\beta$ | *Days* | *Beds* | | | |
| Hosp: | 0.45 | 0.18 | 0.91 | -0.69 | 100.61 | 12 | 54 |  |  |  |
| s.e. | 0.08 | 0.09 | 0.063 | 0.13 | 43.00 |  |  |  |  |  |
| SARIMA(2,1,0)(0,0,1) Box Cox trans: $\lambda=1.2$ $\beta_{t-value}=2.05$ | | | | | | | | | | |
| | $\phi_1$ | $\phi_2$ | $\Theta_1$ | $\beta$ | *Days* | *Beds* | | | | |
| ICU: | 0.12 | 0.29 | 0.15 | 10.18 | 12 | 9 |  |  |  |  |
| s.e. | 0.09 | 0.09 | 0.09 | 4.98 |  |  |  |  |  |  |
| **Veneto** |  |  |  |  |  |  |  |  |  |  |
| SARIMA(2,1,1)(1,0,1) Box Cox trans: $\lambda=1.4$ $\beta_{t-value}=2.07$ | | | | | | | | | | |
| | $\phi_1$ | $\phi_2$ | $\theta_1$ | $\Phi_1$ | $\Theta_1$ | $\beta$ | *Days* | *Beds* | | |
| Hosp: | 0.73 | 0.21 | -0.76 | 0.77 | -0.60 | 341.70 | 13 | 82 |  |  |
| s.e. | 0.11 | 0.09 | 0.08 | 0.15 | 0.19 | 164.82 |  |  |  |  |
| SARIMA(0,1,1)(0,1,1) Box Cox trans: $\lambda=1.34$ $\beta_{t-value}=2.52$ | | | | | | | | | | |
| | $\theta_1$ | $\Theta_1$ | $\beta$ | *Days* | *Beds* | | | | | |
| ICU: | 0.06 | -0.67 | 20.72 | 13 | 12 |  |  |  |  |  |
| s.e. | 0.09 | 0.09 | 8.22 |  |  |  |  |  |  |  |
| **Alto Adige** |  |  |  |  |  |  |  |  |  |  |
| SARIMA(3,1,0)(0,1,1) Box Cox transf: $\lambda=1.69$ $\beta_{t-value}=1.92$ | | | | | | | | | | |
| | $\phi_1$ | $\phi_2$ | $\phi_3$ | $\Theta_1$ | $\beta$ | *Days* | *Beds* | | | |
| Hosp: | 0.27 | -0.25 | 0.36 | -1.00 | 182.07 | 12 | 30 |  |  |  |
| s.e. | 0.08 | 0.08 | 0.09 | 0.07 | 94.66 |  |  |  |  |  |
| SARIMA(0,0,3)(0,1,2)) Box Cox transf: $\lambda=1.98$ $\beta_{t-value}=5.60$ | | | | | | | | | | |
| | $\theta_1$ | $\theta_2$ | $\theta_3$ | $\Theta_1$ | $\Theta_2$ | $\beta$ | *Days* | *Beds* | | |
| ICU: | 1.06 | 1.04 | 0.63 | -0.58 | -0.30 | 27.12 | 12 | 8 |  |  |
| s.e. | 0.06 | 0.07 | 0.06 | 0.13 | 0.10 | 4.84 |  |  |  |  |
| **Piemonte** |  |  |  |  |  |  |  |  |  |  |
| SARIMA(10,1,1)(1,1,1) Box Cox: $\lambda=3$ $\beta_{t-value}=3.82$ | | | | | | | | | | |
| | $\phi_3$ | $\phi_5$ | $\phi_6$ | $\phi_{10}$ | $\Theta_1$ | $\Phi_1$ | $\theta_1$ | $\beta$ | *Days* | *Beds* |
| Hosp: | 0.22 | 0.11 | 0.31 | 0.19 | 0.16 | 0.21 | -1.0 | 211187.31 | 12 | 86 |
| s.e. | 0.08 | 0.07 | 0.08 | 0.07 | 0.09 | 0.09 | 0.05 | 55285.44 |  |  |
| SARIMA(3,1,0)(0,1,1) Box Cox: $\lambda=1.2$ $\beta_{t-value}=2.03$ | | | | | | | | | | |
| | $\phi_1$ | $\phi_2$ | $\phi_3$ | $\theta_1$ | $\beta$ | *Days* | *Beds* | | | |
| ICU: | 0.38 | 0.36 | 0.17 | -0.83 | 60.33 | 12 | 36 |  |  |  |
| s.e. | 0.08 | 0.08 | 0.08 | 0.08 | 29.74 |  |  |  |  |  |

As for the reproduction number it has to be said that, being based only on the known (detected) cases, is not designed to capture the hidden variations generated by the (unknown) asymptomatic. For example, Italy and UK experienced during the summer a strong reduction of the Rt values, which exhibited values below one. However, the data released at the beginning of the month of September, showed that, unfortunately, the virus did not stop spreading in summertime, and the Rt failed to properly react to the ongoing spreading situation. Thus, it is not unreasonable to assume the Rt predictive capacity to be approximately less or equal 4 days, consistently with other available indicators based on officially reported cases. Moreover, it might not be unlikely the reduction of such prediction horizon, considering the computation time actually needed for this indicator to be released.

The impact of under-ascertainment (the ratio of confirmed cases to the true number of cases) on the reproduction number is also discussed in [35], where the correlation between testing and the amount of unknown cases is investigated. In essence, the Rt – Being based on the number of cases officially reported – should be expected to embody biasing components, to an extent directly proportional to the quota of unknown cases.

On the contrary, the TPR, as we have demonstrated, adds an approximate extra time of 11 days (the average number of days between the infection and symptoms onset) leading to a pure predictive time lag of about 15 days, and a “theoretical” one of about 12 days.

Last but not least, TPR precision clearly depends also on the data collection process adopted, which should be designed and implemented to guarantee the lowest possible error rates in the transmission of the test results. This also to minimize the negative impact arising from the above mentioned retrospective revisions. Indeed, it would be possible to define more precise TPR measures provided that the data were organized in a more structured form, as discussed in [19]. It is a fact that, by collecting and making available additional information – often readily available to the health care provider – the TPR would significantly improve its reliability. For example, it might be possible to gain precious insight by simply studying the effectiveness of different test typologies (diagnosis, screening, surveillance for health care operators and so forth) and associating specific accuracy information to the different types of tests administered. Clearly, the more (quality) information enter the TPR the more valuable its contribution in the description and prediction of the covid’s dynamics. For example, data collection could be improved developing point-of-care instant screening tests [39], incorporating TPR data transmission and calculation, as depicted in Figure 4. In this scenario, an improved TPR could be fruitfully exploited for monitoring, surveillance, and forecasting purposes, as well as to integrate electronic health records with information retrieved by sensors[39]. Nevertheless, results obtained in this study emphasize the effectiveness of the proposed approach.

**Figure 4:**
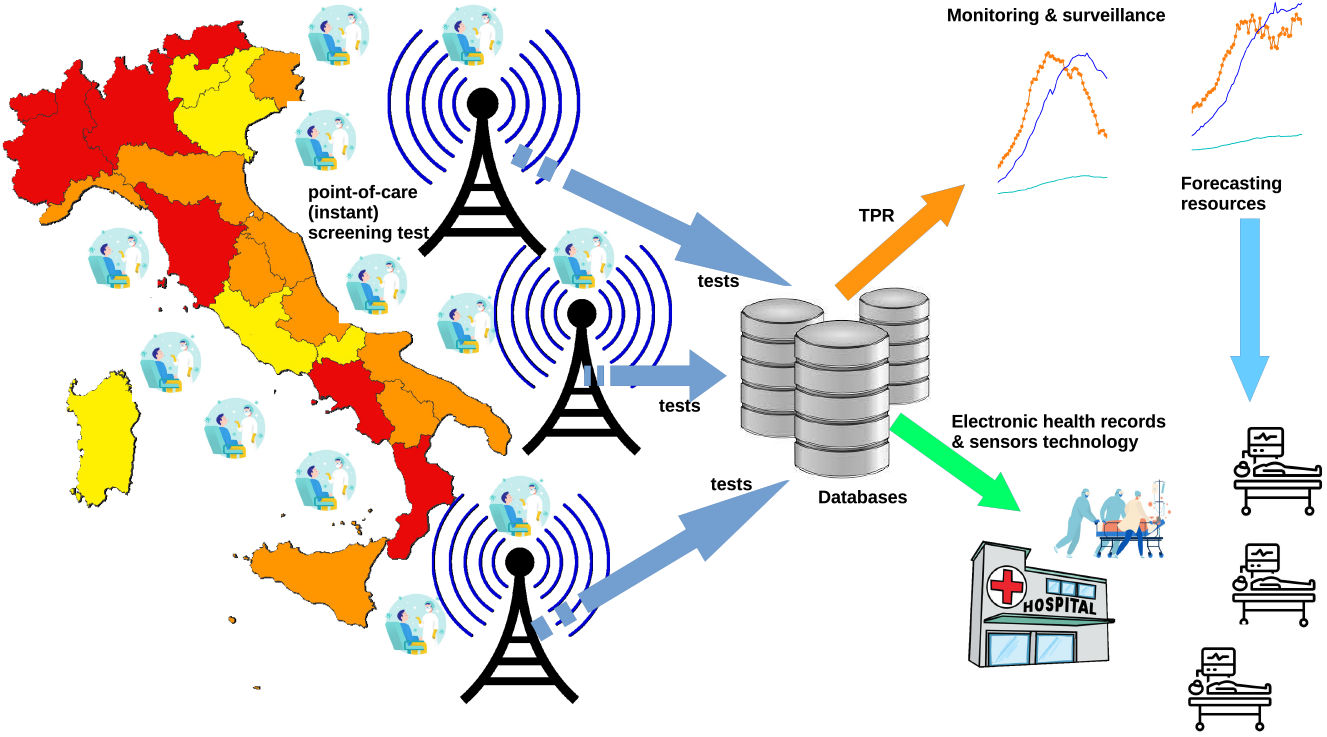
Developing point-of-care (instant) screening tests for COVID-19: data collection, sensors technology, TPR calculation and information flows.

In this paper, we have presented a forecasting method for the short term prediction of the impact of CoViD-19 disease on the public health system. To this end, we have provided enough evidence about the goodness of the TPR as a leading indicator for both the number of people hospitalized and, out of this group, for those who required a bed in intensive care units. The theoretical framework chosen – that is the time series analysis – has been particularly useful for the dynamic comparison and the exploitation of the information contained in the TPR time series. In our simulations, the model chosen, of the type REG-SARIMA, was able to generate reliable predictions from a minimum of 8 to 12 lags. However, especially in light of new developments of the disease – which take the form of many variants – the prediction performances of the REG-SARIMA model might might be affected, if not impaired altogether. Therefore, future directions include the study of a more appropriate model, e.g. of the type regime-switching. Furthermore, additional external information (e.g. the time varying percentage of critical cases) could be fruitfully exploited in a Bayesian theoretical framework (e.g. of the type Bayesian Hidden Markov Models [41]) or using heuristic based approaches (e.g. like the DempsterShafer techniques [5]). Finally, we will consider the remaining Italian regions as soon as time series of “enough” lenght become available.

## 4 Methods

### 4.1 The standardized TPR index

The TPR is one of the metrics commonly used to infer the level of transmission of a disease in a population [8], and, as a such, has been also used in the case of the COVID-19 for different purposes, see for example [20, 30, 33]. However when different types of tests are used, as it happened during the second phase of the pandemic in Italy, where antigen tests have been extensively used, the definition of the TPR becomes more critical. In this study, we will use a standardized version of the TPR index defined by one of the authors [19], which allows to integrate antigen tests in the index calculation.

Following the style of [19], where the Greek letters Θ, Φ and *µ* have been replaced respectively with the letters *τ, ρ* and *ω*, for consistency with the statistic notation later employed, the mean TPR index *τ* on *ω* days is defined as follows:

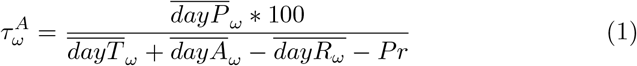

where 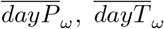 and 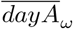 are respectively the average values of new positive cases, molecular (PCR) tests and antigen tests done in the last *ω* days. To compute the TPR index, the average number of healed patients in the last *ω* days, 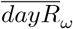 and an estimation for the number of repeated tests *Pr* are subtracted from the total number of tests. We assume that at least one test is done for each healed patient. The number of repeated tests *Pr* is computed using the formula 2, following the approach presented in [19]:

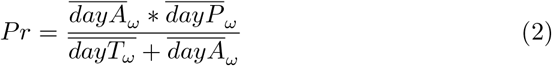

This formula is obtained assumning that the positivity rates for antigen tests and molecular tests are the same, and thus *dayA/Pr* = *dayT/*(*dayP* −*Pr*). Using this approach the computed *Pr* can be considered an upper bound, because the molecular tests positivity rate is generally greater then the one related to antineg tests which are mainly used for screening purposes, see for example [40].

Finally, following the style of [19], a factor *ρ* is added to *τ* in order to model the impact of the number of tests on the remaining susceptible individuals, which are computed removing the total infected cases *I* from the population *N* of a given region. The number of tests are subtracted removing the repeated ones and those used for healed patients, obtaining the following formula:

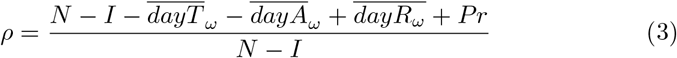

and the TPR index *τ*_*ω*_ is defined as follows:

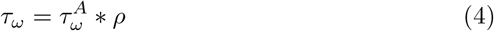

### 4.2 The statistical method applied

Throughout the paper, the time series of interest, say *x*_*t*_, is always intended to be a real–valued, uniformly sampled, sequence of data points of length *T*, formally expressed as

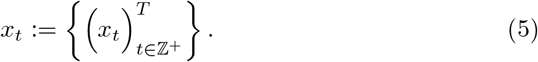

Furthermore, *x*_*t*_ is supposed to be a realization of an underlying stochastic process of the type *SARIMA* (short for Seasonal Auto Regressive Moving Average).

Mathematically, *SARIMA* models take the form of a *t*-indexed difference equation – being *t* as defined in (5) – i.e.:

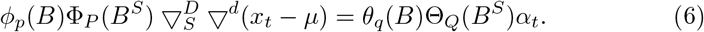

Denoting with *B, d* and *D* the backward shift operator and the non-seasonal and seasonal difference operator respectively, defining ^*d*^ = 1 *B*^*d*^ and ^*D*^ = 1 − *B*^*D*^, we have *φ*_*p*_(*B*) = 1 − *φ*_1_*B φ*−_2_*B*^2^− …. − *φ*_*p*_*B*^*p*^, *θ*_*q*_(*B*) = 1 − *θ*_1_*B* − *θ*_2_*B*^2^ −…. − *θ*_*q*_*B*^*p*^, Φ_*P*_ (*B*^*S*^) = 1 − Φ_1_*B*^*S*^ − Φ_2_*B*^2*S*^− …. − Φ_*P*_ *B*^*P S*^ and Θ_*Q*_(*B*^*S*^) = 1 − Θ_1_*B*^*S*^ − Θ_2_*B*^2*S*^− …. − Θ_*q*_*B*^*QS*^. Here, *φ, θ*, Φ, Θ, respectively denote the non-seasonal autoregressive and moving average parameters and the seasonal autoregressive and moving average parameters. Finally *α*_*t*_ is a 0–mean white noise with finite variance *σ*^2^. In the present paper, external information is exploited and embodied in (6) in the form of a matrix of regressors *D*_*j,t−k*_, with *k* ∈ Z+, weighted by a vector of coefficients *β*_*j*_, i.e.

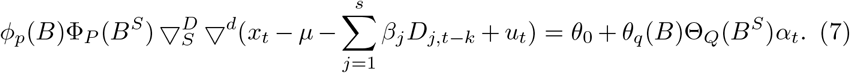

This particular extension is usually referred to as *REG-SARIMA*, to stress the role played by the possibly lagged (of an amount equals to *k* temporal lags) regressors, stored in the matrix *D*_*j,t*_. This types of models are designed to capture the stochastic dynamics generated by the residuals obtained by regressing the matrix *D* (the independent variable(s)) on the time series of interest (the dependent variable). A better insight of the stochastic mechanism governing the *REG-SARIMA* equation can be gained by re-expressing equation 6 so as to emphasize the role played by the term *u*_*t*_ in (7), i.e.

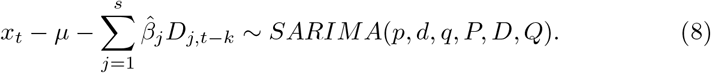

This formulation makes clear the flexibility of this approach which allows the extraction of the significant lags at which the different regressors impact the time series of interest as well as their magnitudes.

If the integration constants *d* and *D* (introduced in Equation 8) are certainly useful to mitigate – if not solve altogether – many stationarity problems, on the other hand they might not be effective against non-normality and/or eteroschedasticity issues. Unfortunately, the data considered in this paper are affected by both these phenomena and therefore, as a coping mechanism, the well-known one–parameter Box–Cox data transformation has been adopted. Presented in the mid-sixties in [6], this method has been discussed and applied in a wide range of problems (see, among others, [36], [22] and [25]), given the widespread acceptance gained over the years. Its mathematical formulation is quite straightforward and takes the form of a power transformation, i.e.

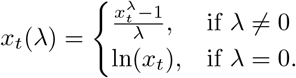

By embodying the *λ* parameter in Equation 7, the model employed in this paper is finally defined, i.e.

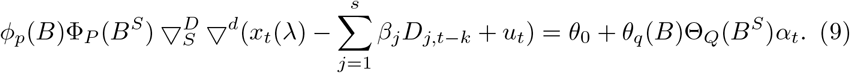

The inference procedures carried out for the estimation of Equation 9 are of two types: maximum likelihood for the *SARIMA* parameters {*φ, θ*, Φ, Θ, *d, D*} and ordinary least squares for the vector *β*. Finally, the hyper-parameters {(*p, d, q, P, D, Q*)} as well as the Box-Cox constant *λ* are estimated within the framework of the Information Theory as explained in the following section.

### 4.3 Estimation of the model order and the *λ* parameter

Akaike’s Information Criterion *AIC* ([1], [9], [21]) – one of the most popular model selector – will be employed to choose the *SARIMA* model order as well as the Box-Cox *λ* parameter. The selection of those constants is not a trivial task as it entails the solution of a conditional multi-objective problem induced by the 6–dimensional vector of unknown constants Γ ≡ {(*p, d, q, P, D, Q*)} conditional to the Box-Cox paramter *λ*. The estimation method employed to find the “best” conditioned vector of hyper-parameters – that is the one governing the selected order structure 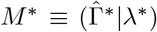 – relies on the information theory and, in particular, on the Akaike Information Criterion (AIC). At its core, *AIC* is based on an estimate of the expected relative entropy (the Kullback–Leibler divergence) contained in an estimated model, that is the degree of divergence from the “true” theoretical model. Assuming *X*_*t*_ to be randomly drawn from an unknown distribution *H*(*x*), with density *h*(*x*), estimation of *h* is done by means of a parametric family of distributions with densities [*f* (*x*|*θ*; *θ* ∈ Θ)], *θ* the unknown parameters’ vector. Denoting by 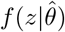 the predictive density function, by *f* the true model and by *h* the approximating one, Kullback-Leiber divergence takes the form

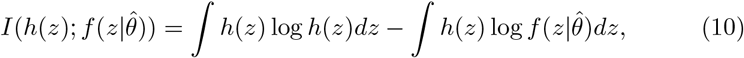

which, after some algebra, can be written as follows:

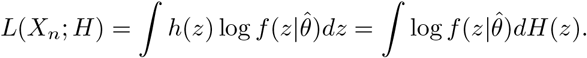

This quantity can be estimated by replacing *H* with its empirical distribution *Ĥ*, so that 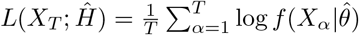. This is an overestimated quantity of the expected log likelihood, given that *Ĥ* is closer to 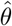 than *H*. The related bias can be written as follows:

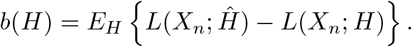

Denoting, by the Greek letter *ξ* the number of estimated parameters, Akaike proved that 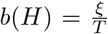, so that the information based criterion takes the form 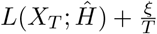. By multiplying this quantity by −2, finally *AIC* is defined as

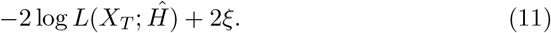

Elaborating on [34], the correct formulation of AIC for the model expressed in Equation 9 takes the form

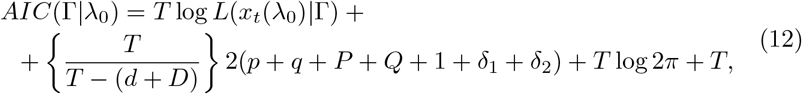

where

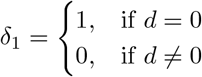

and

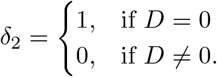

By sequentially applying Equation 11 for different combinations of the hyper-parameters {(*p, d, q, P, D, Q*))} and conditioning the observed data to *a given λ* parameter (which in Equation 12 has been denoted with *λ*_0_) a sequence of AIC values is obtained. This is the first of the two-step selection strategy adopted in the present paper, which is usually referred to as *MAICE* (short for Minimum *AIC* Expectation) [2] procedure. In the second step, the order (Γ^*\**^) satisfying:

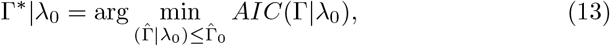

i.e. the minimizer of the AICs generated by the candidate models, will be the winner model structure. However, Equations 12 and 13 are not designed to estimate the Box-Cox *λ* parameter. To this end, a grid search approach – over a set Λ of *B* competing parameters {*λ*_*j*_; *j* = 1, 2, …, *B}* – has been applied. Each *λ* has been evaluated in terms of the contributions given in terms of both data normalization and statistical significance of the external regressor. Finally, *MAICE* procedure requires the definition of an upper bound for all the Γ parameters, as a maximum order a given process can reach. This choice, unfortunately, is *a priori* and arbitrary.

## Data Availability

The TPR data for all Italian region compared with hospitalized time series is available in the following Web page.

http://www.cs.unibo.it/~gaspar/www/italy.html

## Acknowledgements

The author would like to thank the Italian Civil Protection Department, and all the staff involved for providing the data of the outbreak used in this study.

## Author Contributions

- **Livio Fenga:** his contribution concerns the development of the statistical method used for the lagged correlation analysis and the construction of the prediction model. He also wrote the article and contributed to the discussion of the results.
- **Mauro Gaspari:** his contribution concerns the definition of the TPR index and the design of the scenarios as well as of the associated figures. He also designed the forecasting experiments, prepared the time series. He finally wrote the article and contributed to the discussion of the results.

## Footnotes

1 In the three-tiered system issued in Italy to combat the spread of COVID-19, the “red zone” indicates an high-contagion-risk area where non-essential shops and markets are closed and residents are only allowed to leave their homes for work, health reasons or emergencies.

